# The impact of COVID-19 on the implementation of routine immunisation for children in Rwanda

**DOI:** 10.1101/2025.04.09.25325536

**Authors:** Edward Mbonigaba, Fengyun Yu, Mark Donald C. Reñosa, Frederick Nchang Cho, Qiushi Chen, Wenjin Chen, Claudia M. Denkinger, Shannon A. McMahon, Simiao Chen

**Author notes:** Correspondence should be addressed to Mbonigaba Edward.

## Abstract

**Introduction:** The most common indirect impact of the Coronavirus Disease 2019 (COVID-19) pandemic has been the interruption of routine immunisation services, resulting in a significant decline in childhood immunisation rates, particularly during the early stages of the pandemic. This study aimed to explore the socio-demographic characteristics associated with continued routine immunisation in Rwanda.

**Methods:** A cross-sectional survey was conducted between January 3^rd^ to March 31^st^, 2022 amongst mothers from five districts in Rwanda. Multinomial logistic regression was used to determine associations between demographic characteristics and the willingness to vaccinate children, considering the impact of the COVID-19 pandemic on vaccination attitudes.

**Results:** Among the 2,045 mothers surveyed, 92.2% and 91.6% admitted that their religion and culture support immunisation respectively. Marital status, educational status, and average monthly income were significantly associated with culture and tradition. Out of the 2,045 mothers, 77.3% and 58.7% were concerned about the serious adverse effects of vaccines and COVID-19 vaccines, while 8.1% were concerned with the safety of COVID-19 vaccines. With the exception of age, marital status, and the number of children in the immunisation age bracket, there was a significant association between the perceived risks of vaccination and all other socio-demographic characteristics.

**Conclusion:** Global routine immunisation was disrupted throughout the duration of the COVID-19 pandemic. However, Rwanda’s initial preparedness to combat infectious diseases such as Ebola minimized the influence of COVID-19 on routine immunisation in the country. This study suggests an association between routine immunisation and factors such as culture, financial constraints, vaccine misinformation, concern about adverse effects of vaccines, and apprehensions over the safety of COVID-19 vaccines.

## Introduction

The spread of the new Coronavirus Disease 2019 (COVID-19), caused by the virus known as Severe Acute Respiratory Syndrome Coronavirus (SARS-CoV-2), originated in Wuhan, China in December 2019[1–3]. By 2021, the emerging variants of COVID-19 have affected around 230 million individuals globally, leading to 4.7 million deaths[4]. The number of infections surpassed 100 million people in 2020[5,6]. In Africa, there were 12.4 million cases and 256,000 deaths[7], with Rwanda recording 133,194 cases and 1,468 deaths[8].

One of the most common indirect impacts of the COVID-19 pandemic was the disruption of routine immunisation (RI) services[9,10]. In Rwanda, there was a significant increase in visits to hospital by 60%, along with a corresponding decrease in visits to health centres by 15% [11,12]. This disruption was not limited to Rwanda; globally, around 23 million children worldwide missed basic immunisations, 30 million missed the recommended doses of diphtheria, tetanus and pertussis (DPT3), and 27.2 million children missed the recommended doses of measles-containing-vaccine (MCV1)[10,13–15].

In response to the pandemic, various countries adopted strategies based on their socio-demographic, cultural and political commitments[16] to maintain the continuity of routine immunisation. In March 2021, Rwanda received approximately 240,000 doses of the Oxford AstraZeneca COVID-19 vaccine, and 103,000 doses of Pfizer-BioNTech COVID-19 vaccines through the COVAX Facility platform, supported by organisations such as the Coalition for Epidemic Preparedness Innovations (CEPI), Gavi, the Vaccine Alliance, and World Health Organisation (WHO), in partnership with the United Nations Children’s Fund (UNICEF)[17–19]. The country also received a second batch of these vaccines from India in the same month of 2021[20]. Other COVID-19 vaccines used to mitigate the pandemic were Johnson & Johnson, Moderna, Sinopharm, and Sputnik V[21,22]. The completion of multiple doses of COVID-19 vaccines is essential for ensure a robust immune response, with vaccines like Johnson & Johnson, Oxford-AstraZeneca and Moderna requiring two doses, while for Pfizer-BioNTech may require two or three doses [18,19,23,24]. Achieving a high rate of vaccination uptake and ensuring completion of the recommended doses are critical steps in mitigating the COVID-19 pandemic[25–27] and preventing other vaccine-preventable diseases. The COVID-19 pandemic brought about widespread global disruption in routine childhood immunisation especially in the initial pandemic phases[9,10], especially with the lockdowns.

The pandemic and subsequent vaccine rollout created significant challenges with vaccine hesitancy (VH). Many countries including Rwanda, faced obstacles in ensuring high uptake and completion of the COVID-19 vaccination schedule. Vaccine hesitancy is influenced by various factors such as misinformation, perceived risks, and concerns about the safety of vaccines. Studies highlighted the role of anti-vaccine movements[28] and accessibility[22] issues, which impacted the continuation and completion of RI. Additionally, concerns about side effects and safety of COVID-19 vaccines further exercebated the challenge of achieving widespread vaccine coverage[23,29,30].

While global research has focused on vaccine acceptance, hesitance, and attitudes towards COVID-19 vaccines, there is limited literature specific to the continuation of RI in the context of the pandemic, particularly in Rwanda. Existing studies predorminantly concentrate on vaccine uptake for COVID-19 vaccines, but few explore the broader impact on routine childhood immunisation during the pandemic [31–33]. This gap is evident and leaves a critical need for more context-specific research, especially in countries like Rwanda, where socio-demographic factors such as culture, financial constraints, and vaccine misinformation may significantly influence immunisation behaviours [34,35].

The aim of this study is to fill the gap by exploring the socio-demographic characteristics associated with the continuation of RI in Rwanda during the COVID-19 pandemic.

## Materials and Methods

### Study design and participants

A national cross-sectional study was conducted between January 03^rd^ to March 31^st^, 2022, across five districts in Rwanda: Nyagatare and Ngoma in the Eastern Province, and Nyarugenge, Nyamagabe, and Ngororero in the Central, Southern, and Western Provinces respectively.

Rwanda is a country in the Great Lakes region of Africa, covering 26,338 square kilometres, with a population of 14.14 million people in 2021 and a growth rate of 2.38% between 2019 and 2020 [36,37]. The median age of the Rwandan population is 9.4 years[38], and the capital city, Kigali has a population of 745,261 people [37]. Although a low-income country, with over 70% of the population engaged in agriculture, Rwanda aspires to reach middle-income status by 2035 and high-income status by 2050[37,39].

Study participants were mothers aged ≥ 18 years who had children 12 – 23 months, spoke English, and provided written informed consent to participate in the study. Mothers who did not have children in the immunisation age group, male participants, and caregivers other than mothers were excluded. Data were collected from 50 villages in the selected districts.

### Sampling method and sample size calculation

A multi-stage cluster sampling method was employed. First, the five provinces were listed, and four provinces (Easter, Central, Southern, and Western) were randomly selected. One distict was randomly selected from each province, and within each district, two sectors were selected. For each sector, two “Akagalis” (villages) were chosen. A total of 50 villages in five health districts of Rwanda were chosen for data collection (Figure 1).

**Figure 1:**
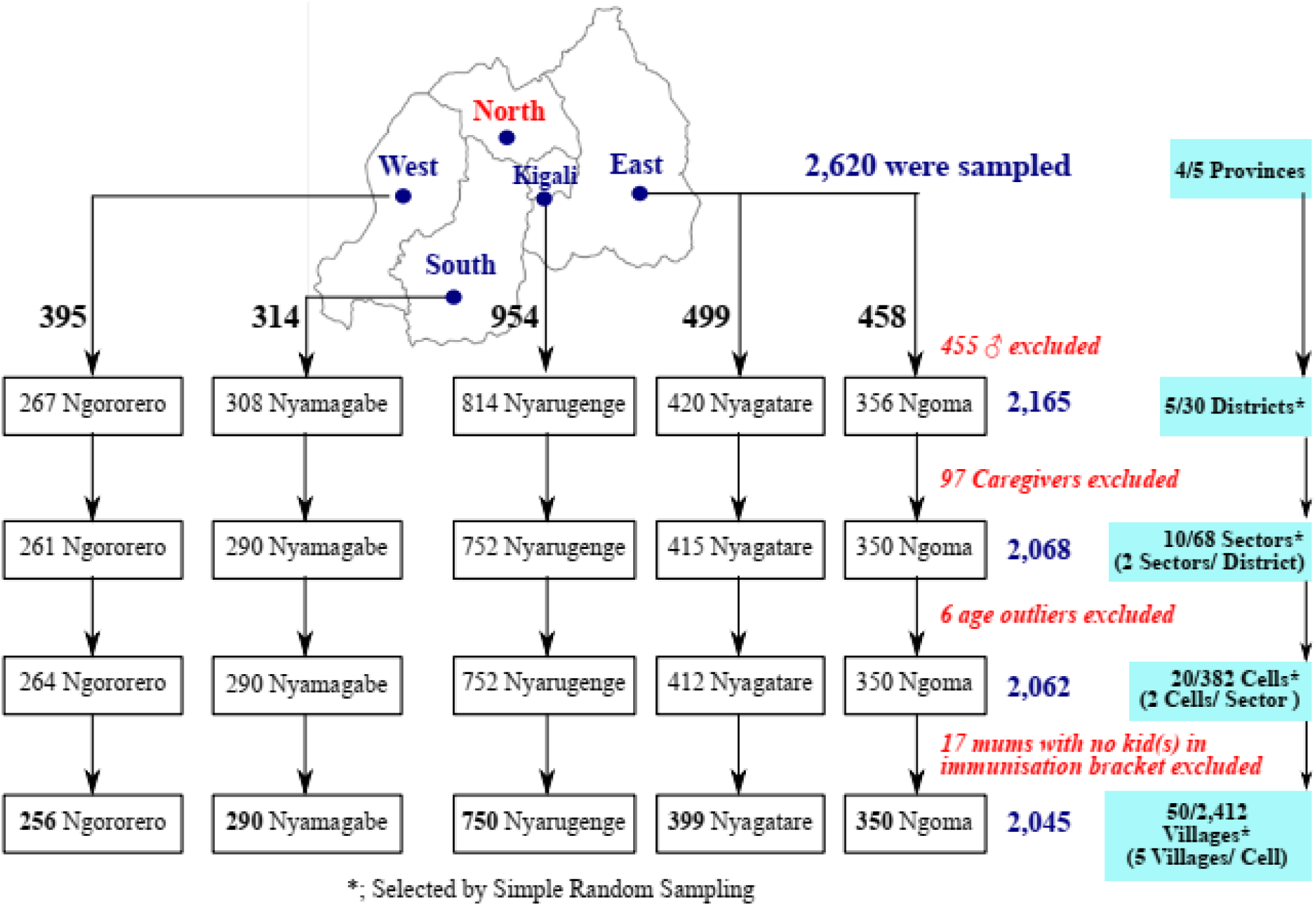
Schematic illustration of the study design, settings, and eligibility of respondents.

In each selected village, mothers were consecutively and conveniently recruited intil the desired sample size was achieved. In the absence of similar studies in Rwanda, a minimum sample size of 154 mothers per Cluster/District, based on the WHO immunisation coverage cluster survey[40], was calculated with the CDC Epi Info version 7.2.5.0 (Centre for Disease Control, Georgia, USA) StatCalc with the following characteristics: an estimated District population size of 362,806 in 2022[37], an estimated proportion of mothers who continued routine immunisation after the emergence of the COVID-19 pandemic of 50.0%, a design effect of 2.0, an accepted error margin of 5%, and five Districts. Considering possible non-response and non-responding respondents, the sample size was adjusted by 10% (16 respondents) to 170.

### Definition of concepts and study variables

#### Independent and demographic variables

Socio-demographic variables included age, religion, marital and educational status, number of children in the immunisation bracket, occupation, and average monthly income. Other independent variables were religion and culture (Q23–24; S1 Appendix), and lack of funds (Q25; S1 Appendix).

#### Outcome variables

The primary outcome was the effect of the COVID-19 pandemic on the willingness to get children vaccinated. Other outcome variables included the effect of religion, culture, economic status and perception of risk on the children’s routine vaccination and the degree of risk perception, misinformation and trust. The questions and coding schemes of the above outcome variables are outlined in Table 1.

**Table 1:** Question for outcome variables.

| Index | Category | Question | Response(s) | Coding |
| --- | --- | --- | --- | --- |
| XX | Primary outcome | Has the COVID-19 pandemic affected your routine immunisation of your children? | - Yes;<br>- No | - Yes = 0;<br>- No = 1 |
| Q23 | Religion | Does your religion support the Immunisation of children? | - Yes;<br>- No | - Yes = 0;<br>- No = 1 |
| Q24 | Culture | Does your culture support the Immunisation of children? | - Yes;<br>- No | - Yes = 0;<br>- No = 1 |
| Q25 | Economic status | Does lack of funds ever prevent you from Immunisation of your child? | - Yes;<br>- No;<br>- I don't know | - Yes = 1;<br>- No = 0;<br>- I don't know 1 |
| Q26 | Perception of risk | Does the message you hear or receive from anti-vaccine groups or propaganda affect your confidence in the vaccination of your children? | - Yes;<br>- No | - Yes = 0;<br>- No = 1 |
| Q27 | Trust/Confidence | Do you trust the information you receive about the vaccination of your children? | - Yes;<br>- No | - Yes = 0;<br>- No = 1 |
| Q38 | Perception of risk | I am concerned about the serious adverse effects of vaccines. | - Strongly Disagree,<br>- Disagree,<br>- Undecided/Neutral,<br>- Agree,<br>- Strongly Agree | - **Disagree**; Strongly Disagree, Disagree, Undecided/Neutral, **Agree**; Agree, Strongly Agree |
| Q50 | Perception of risk | I am concerned about the serious adverse effects of COVID-19 vaccines | - Strongly Disagree,<br>- Disagree,<br>- Undecided/Neutral,<br>- Agree,<br>- Strongly Agree | - **Disagree**; Strongly Disagree, Disagree, Undecided/Neutral, **Agree**; Agree, Strongly Agree |
| Q53 | Perception of risk | I am concerned that COVID-19 vaccines might not be safe. | - Strongly Disagree,<br>- Disagree,<br>- Undecided/Neutral,<br>- Agree,<br>- Strongly Agree | - **Disagree**; Strongly Disagree, Disagree, Undecided/Neutral, **Agree**; Agree, Strongly Agree |
| Q56 | Misinformation | Received negative information about COVID-19 vaccines | - Yes;<br>- No | - Yes = 0;<br>- No = 1 |
| Q57 | Trust/Confidence | The trust in negative information on the COVID-19 vaccine | - Yes;<br>- No | - Yes = 0;<br>- No = 1 |

VA; Vaccine Acceptance, VH; Vaccine Hesitancy

The potential responses to each of these questions were sorted on a Likert scale[41] and included Strongly disagree, Disagree, Undecided, Agree, and Strongly agree. These potential responses were then scored as one, two, three, four, and five, respectively. Modified Bloom’s cut-off points were used to rate the perception of risk of vaccination as very poor (< 20%), poor (≥ 20 but < 40%), moderate (≥ 40 but < 60%), good (≥ 60 but < 80%), or very good (≥ 80%) and later as poor (< 60%) or good (≥ 60%)[42,43].

### Sampling method

We implored a multistage Cluster (province) sampling method where a list of all Cells/“Akagalis” and villages therein was drawn. A total of 50 villages were selected, including at least two villages from each Cell/“Akagali”. The sampling procedure for the required number of mothers was done in three stages.

Firstly, the five provinces were listed and then four; the Western, Southern, Centre/Kigali, and Eastern provinces were selected by using random proportions of the RAN function in Microsoft Office Excel, 2016 (Microsoft Corporation Inc. USA), wherein the least proportion attributed to the Northern province was not selected. Secondly, the number of districts of the selected provinces was listed, and one of each was selected using Simple Random Sampling (SRS). Within each district, two sectors were selected and within each sector, two Akagalis were selected. At least two villages were then selected from each Akagali by SRS (Figure 1). Thirdly, within each selected village, mothers were sampled conveniently and consecutively until the desired sample size was attained or surpassed. The convenience and consecutive sampling techniques were applied wherein participants were approached and informed of the study objectives at their workplaces and doorsteps. Data was then collected through personal interviews.

### Data collection and statistical analysis

Data was collected using well-structured questionnaires in a face-to-face interviews which to 20 – 30 minutes to complete. The questionnaire, prepared in Englishe was aimed to collect information on respondents’ identification, demographic characteristics (age, religion, marital status, occupation, average monthly income), information about child(ren) immunisation status, RI, the perception of risks and others. The validity of the questionnaire was confirmed by pre-testing in 10 participants who were excluded from the analysis. Based on the pre-test study, the format and wording of some questions were refined. The data obtained from the 10 participants was used to assess internal consistency reliability using Cronbach’s alpha (α)[44–46]. The results showed adequate internal consistency reliability (with Cronbach’s α = 0.72)[45,46] for the eight sections with 62 questions.

At the end of each day or after every two days of field data collection, the data quality control officers checked the entry of all data into the Microsoft Office Excel sheet to ensure that the right data was being collected. Data was reviewed and sorted multiple times to removed duplicated data before exported to Microsoft Office Excel sheet. At the end of the data collection exercise, the field supervisor checked all the data from the various districts to ensure that the data collected was in order.

### Covariates and outcomes

Age groups, religion, marital status, educational status, number of child(ren) in the immunisation bracket, occupation and average monthly income group were summarised as counts and percentages. Multivariate/multinomial logistic regression was also used to determine associations between the outcome variables (effect of COVID-19 on routine immunisation of children/dependent variables with demographic characteristics.

Multicollinearity was tested for, and the following models were used:

Culture supports the immunisation of children = β_0_ + β_1_Age Group + β_2_Religion + β_3_Marrital Status + β_4_Education + β_5_Number of children in the immunisation bracket + β_6_Occupation + β_7_Monthly Income + ε,

Religion supports the immunisation of children = β_0_ + β_1_Age Group + β_2_Religion + β_3_Marrital Status + β_4_Education + β_5_Number of children in the immunisation bracket + β_6_Monthly Income + ε.

Good perception of vaccine risks = β_0_ + β_1_Age Group + β_2_Religion + β_3_Marrital Status + β_4_Education + β_5_Number of children in the immunisation bracket + β_6_Occupation + β_7_Monthly Income + ε, and

Anti-vaccine misinformation/Trust of misinformation = β_0_ + β_1_Age Group + β_2_Religion + β_3_Marrital Status + β_4_Education + β_5_Number of children in the immunisation bracket + β_6_Occupation + β_7_Monthly Income + ε. and.

Effects of COVID-19 on RI = β0 + β1Culture + β2Lack of funds + β3Vaccine misinformation + β4Trust of vaccine misinformation + β5Concerned about AEs of vaccines + β6Concerned about AEs of COVID-19 vaccines + β7Concerned that COVID-19 vaccines might not be safe + ε.

Where β_0_ is a constant, β_1_, β_2_, β_3_, β_4_, β_5_, β_6_, and β_7_ are coefficients and ε is the regression error. For multicollinearity, a variance inflation factor between 1 and 5 indicated a moderate correlation between a given predictor variable and other predictor variables in the model [47]. The significance level was set at 0.05. Data were analysed using CDC Epi Info version 7.2.5.0 (Centre for Disease Control, Georgia, USA).

### Ethical consideration

This study was conducted in strict accordance with the Declaration of Helsinki[48] and approved by the Institutional Review Board (IRB) of the University of Rwanda, College of Medicine and Health Science (No. 402/CMHS IRB/2020) and the Ethics Committee of Universität Heidelberg, Germany (S-829/2021). All study participants signed the informed consent before being interviewed. All participants were informed and assured that the data collected would be used only for research purposes and that their responses would not be available to the public.

## Results

### Characteristics of Study Population

Socio-demographic characteristics are summarised in Table 2. A total of 2,620 respondents were surveyed, from which 2,045 (78.2%) mothers were included in these analyses as follows: 350 (17.1%) from Ngoma, 256 (12.5%) from Ngororero, 399 (19.5%) from Nyagatare, 290 (14.2%) from Nyamagabe, and 750 (36.7%) from Nyarugenge.

**Table 2:** Socio-demographic characteristics of study participants (*n* = 2,045)

| Variable | Subclass | District | | | | | | | $\chi^2$ (F) | p-value |
| --- | --- | --- | --- | --- | --- | --- | --- | --- | --- | --- |
|  |  | Total (%) | Percent (95% C.I) | Ngoma | Ngororero | Nyagatare | Nyamagabe | Nyarugenge |  |  |
| Age (in years) | ≤ 30 | 1,072 (52.4) | 52.4 (50.3 - 54.6) | 186 | 116 | 253 | 151 | 383 | 25.835 | **1.1x10<sup>-4</sup>** |
|  | 31 – 40 | 874 (42.7) | 42.7 (40.6 - 44.9) | 139 | 119 | 149 | 131 | 336 |  |  |
|  | ≥ 41 | 99 (4.8) | 4.8 (4.0 – 5.9) | 45 | 21 | 15 | 8 | 30 |  |  |
| | Mean age ( $\bar{x} \pm$ SD) | 30.6 ± 6.3 | | 30.3 ± 6.6 | 31.7 ± 6.5 | 29.4 ± 6.2 | 30.6 ± 5.8 | 30.9 ± 6.1 | 6.666 | **<0.001** |
| Religion | Catholic | 85 (4.2) | 4.2 (3.4 - 5.1) | 6 | 1 | 8 | 22 | 48 | 30.029 | **<0.001** |
|  | Protestant | 1,960 (95.8) | 95.8 (94.9 - 96.6) | 344 | 255 | 391 | 268 | 702 |  |  |
| Marital status | Divorced | 59 (2.9) | 2.9 (2.2 - 3.7) | 5 | 1 | 11 | 10 | 32 | 31.296 | **1.8x10<sup>-3</sup>** |
|  | Married | 1,586 (77.6) | 77.6 (75.7 - 79.3) | 269 | 199 | 315 | 213 | 590 |  |  |
|  | Single | 373 (18.2) | 18.2 (16.6 - 20.0) | 71 | 52 | 71 | 57 | 122 |  |  |
|  | Widowed | 27 (1.3) | 1.3 (0.9 - 1.9) | 5 | 4 | 2 | 10 | 6 |  |  |
| Education | NFE | 190 (9.3) | 9.3 (8.1 – 10.6) | 48 | 16 | 45 | 63 | 18 | 522.405 | **<0.001** |
|  | Primary | 963 (47.1) | 47.1 (44.1 – 49.3) | 251 | 192 | 218 | 106 | 196 |  |  |
|  | Secondary | 809 (39.6) | 39.6 (37.5 - 41.7) | 48 | 45 | 130 | 112 | 474 |  |  |
|  | Tertiary | 83 (4.1) | 4.1 (3.3 – 5.0) | 3 | 3 | 6 | 9 | 62 |  |  |
| # of child(ren) | 1 | 1,969 (96.3) | 96.3 (95.4 - 97.0) | 335 | 246 | 380 | 286 | 722 | 5.989 | 2x10<sup>-1</sup> |
|  | 2 – 4 | 76 (3.7) | 3.7 (2.9 - 4.6) | 15 | 10 | 19 | 4 | 28 |  |  |
| | Mean child(ren) ( $\bar{x} \pm$ SD) | 1.04 ± 0.2 | | 1.05 ± 0.3 | 1.06 ± 0.3 | 1.05 ± 0.2 | 1.03 ± 0.3 | 1.04 ± 0.2 | 0.772 | 5.4x10<sup>-1</sup> |
| Occupation | Artisan | 1,143 (55.9) | 55.9 (53.7 - 58.0) | 332 | 212 | 258 | 265 | 76 | 1,133.841 | **<0.001** |
|  | Casual labour | 192 (9.4) | 9.4 (8.2 - 10.7) | 4 | 11 | 13 | 22 | 142 |  |  |
|  | Civil Servant | 110 (5.4) | 5.4 (4.5 - 6.4) | 2 | 14 | 30 | 0 | 64 |  |  |
|  | Unemployed | 600 (29.3) | 29.3 (27.4 – 31.4) | 12 | 19 | 98 | 3 | 468 |  |  |
| AMI (in \$) | < 100 | 1,538 (75.2) | 75.2 (73.3 - 77.0) | 240 | 255 | 356 | 273 | 414 | 396.124 | **<0.001** |
|  | 101 – 200 | 193 (9.4) | 9.4 (8.3 – 10.8) | 43 | 1 | 36 | 17 | 96 |  |  |
|  | 201 – 300 | 167 (8.2) | 8.2 (7.1 - 9.4) | 37 | 0 | 6 | 0 | 127 |  |  |
|  | > 400 | 147 (7.2) | 7.2 (6.2 – 8.4) | 30 | 0 | 1 | 0 | 116 |  |  |
| Total (%) |  | 2,045 | 100.0 | 350 (17.1) | 256 (12.5) | 399 (19.5) | 290 (14.2) | 750 (36.7) |  |  |

1\$ = 1.161 RWF = 0.912 €[49], #; number, %; proportion of respondents, $\bar{x}$ ; Mean, 95% C.I; 95% Confidence interval, AMI; Average Monthly Income, **Boldface** numbers indicate significant *p* values, F; F-Statistic (Fisher test), NFE; No Formal Education, SD; Standard Deviation.

Amongst these participants, [1,072 (52.4%, 95%C.I; 50.3 – 54.6)] were 30 years or less [mean age of 30.6 years (SD 6.3, range 18 – 56)], about two-fifths [809 (39.6%, 95%C.I; 37.5 – 41.7)] had attained a secondary level of education, about three-quarters [1,538 (75.2%, 95%C.I; 73.3 – 77.0)] earned less than one United States Dollar (USD) per month, and about three-quarters [1,586 (77.6%, 95%C.I; 75.7 – 79.3)] were married (Table 2).

Bivariate analysis revealed that, except for the number of child(ren) in the immunisation bracket, all the other characteristics of the study participants were associated with the Districts (Table 2).

### Sources of child vaccination information

A large majority of the mothers (94.1%, 95%C.I; 93.0 – 95.1) had vaccine information from healthcare workers (Medical Doctors and Nurses), about a quarter (28.8%, 95%C.I; 26.9 – 30.8) from the Mass media (Radio and Television), and a very minute (7.1%, 95%C.I; 6.1 – 8.3) proportion from relationships (friends, co-workers, and neighbours) (Supplementary Table 1).

### Reasons for vaccination/immunisation discontinuation

A total of 1,781 participants (96.6%, 95% C.I; 95.6 – 97.3) reported that they had no problem with taking their children for vaccination. Among the participants who did not take their children for vaccinations, only 122 (5.5%; 95% C.I; 4.6 - 6.6) cited inaccessibility to healthcare facilities and 44 (2%, 95% C.I; 1.5 - 2.7) indicated that they do not believe that vaccines are good for their children as reasons.

On the anticipated most important reasons for the non-completion of vaccination doses, 99 (4.8%, 95% C.I; 4.0 – 5.8) of the mothers asserted that the child was healthy, 99 (4.8%, 95% C.I; 4.0 – 5.8) were sceptical by indicating that they were not sure the vaccines were good for their children, while four (0.2%, 95% C.I; 0.08 – 0.5) stated that neighbours children suffered complications from vaccination shots (Supplementary Table 2).

In a separate question, the mothers were asked; ‘Does lack of funds ever prevented you from immunisation of your child’, and only 198 (9.7%, 95% C.I; 8.5 – 11.0) of them said ‘yes’. A hundred and forty-eight (74.7%) of the 198 were artisan workers, and 185 (93.4%) earned less than $100 in a month. The lack of funds or transport to the health facility was significantly associated with the occupation and average monthly income (Supplementary Table 3).

### Religious and traditional tendencies towards the immunisation of children

Of the 2,045 mothers, 1,886 (92.2%, 95% C.I; 91.0 – 93.3) and 1,873 (91.6%, 95% C.I; 90.3 –92.7) admitted respectively that their religion and their culture supported the immunisation of children.

Logistic regression analysis revealed that marital status, educational status, and monthly income were significantly associated with, ‘Does religion’ and ‘Does culture’ support the immunisation of children? From the regression analysis, the odds for religion vs culture to support the immunisation of children was higher amongst divorcees (*p* = 4.5×10^−1^, aOR; 1.2, 95%C.I; 0.7 – 2.3) vs (*p* = 7.3×10^−1^, aOR; 1.1, 95%C.I; 0.6 – 1.9) when compared with married women, singles, and widows (Table 3).

**Table 3:**
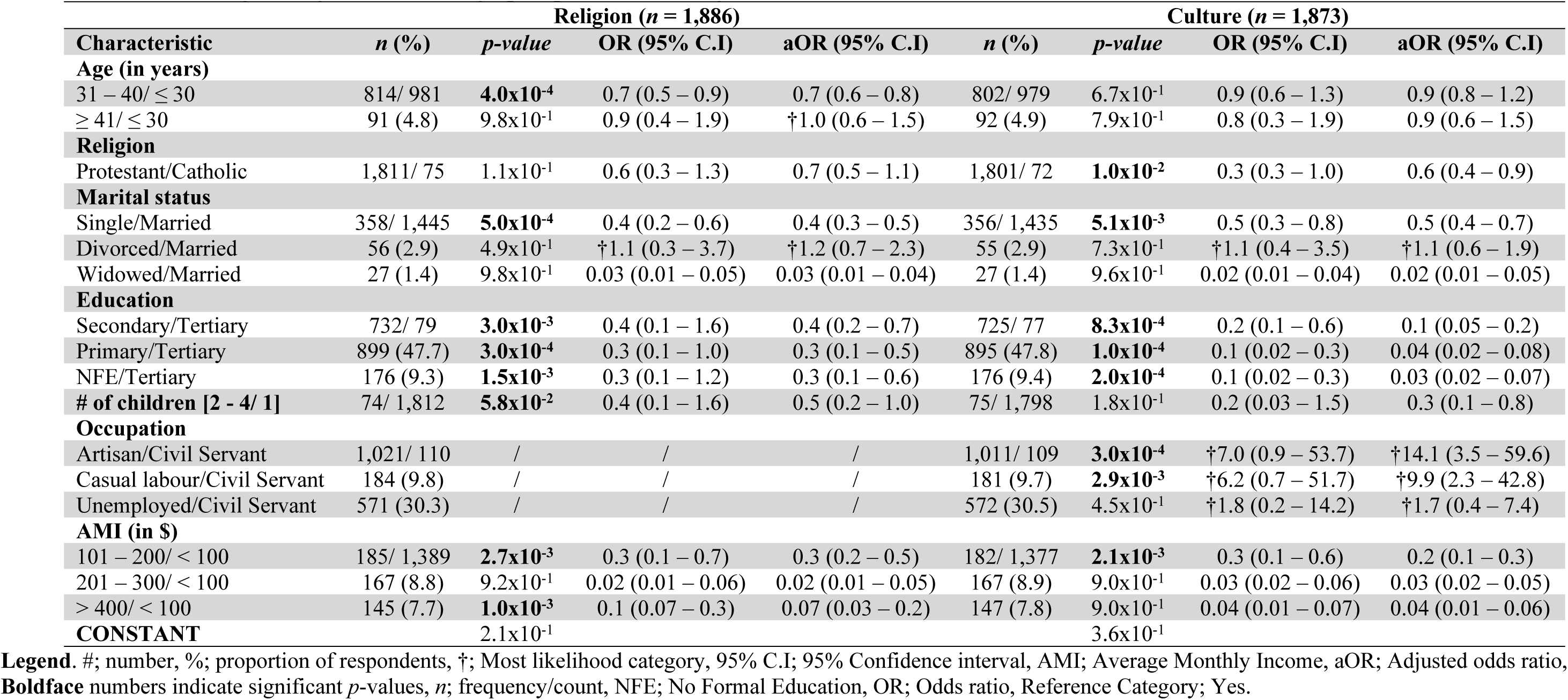
Multivariable logistic regression of demographic predictors of religious and traditional tendencies towards routine immunisation (RI)

### Mothers’ perception of risks

Of the 2,045 mothers in the study, 1,580 (77.3%, 95% C.I; 75.3 – 79.0), 1,201 (58.7%, 95% C.I; 56.6 – 60.8), and 166 (8.1%, 95% C.I; 7.0 – 9.4) of the mothers agreed to the fact that; there were serious adverse effects (AEs) of vaccines, serious AEs of COVID-19 vaccines, and that the COVID-19 vaccines may not be safe respectively. Less than one-quarter [465 (22.7%, 95% C.I: 21.0 – 24.6)] of the respondents disagreed on the existence of serious AEs of vaccines, while 844 (41.3%, 95% C.I; 39.2 – 43.4) of them disagreed the existence of serious AEs of COVID-19 vaccines as well as a large majority [1,879 (91.9%, 95% C.I: 90.6 – 93.0)] disagreed on the fact that COVID-19 vaccines may have safety concerns (Figure 2).

**Figure 2:**
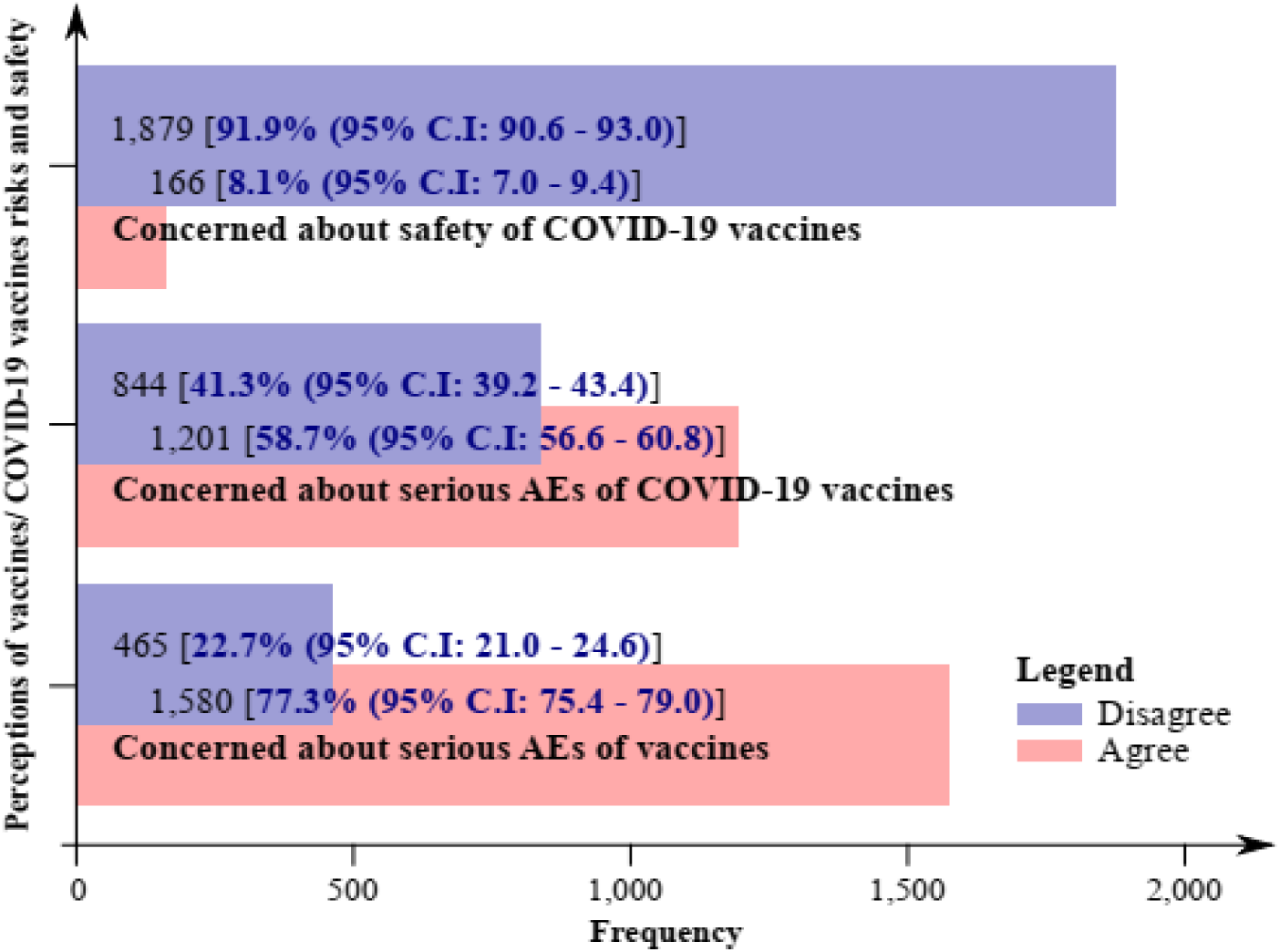
Perceptions of vaccines/COVID-19 vaccines risks and safety.

The perception of vaccine risks and safety by respondents was obtained from the summing up together of; ‘I am concerned about the serious AEs of vaccines’, ‘‘I am concerned about the serious AEs of COVID-19 vaccines’, and ‘I am concerned about the safety of COVID-19 vaccines’. Thus, for the perception of vaccine risks, 1,276 (62.4%, 95% C.I; 60.3 – 64.5) had a good perception; mean vaccine perception score of 61.2 (SD 14.1, range 20.0 – 100.0).

Bivariate and multivariate logistic regression analyses revealed that, except age, and marital status, all the socio-demographic characteristics were significantly associated with the perceived risks of vaccination. From the multivariate regression analysis, the odds for having a good risk perception of vaccines were higher amongst mothers of ≥ 41 years (*p* = 2.1×10^−1^, OR; 1.2, 95%C.I; 0.8 – 1.7), protestants (*p* = 1.7×10^−2^, aOR; 2.5, 95%C.I; 1.6 – 3.9), divorcees (*p* = 9.3×10^−1^, aOR; 1.1, 95%C.I; 0.7 – 1.5), primary school leavers (*p* = 6.3×10^−3^, aOR; 2.1, 95%C.I; 1.2 – 3.6) as well as those with NFE (*p* = 1.0×10^−4^, aOR; 5.1, 95%C.I; 2.8 – 8.9), when compared with their various counterparts; mothers between 31 – 40 years old, Catholic Christians, married mothers and secondary vs tertiary school leavers (Table 4).

**Table 4:** Perception of vaccine risks.

| S/N | Characteristic | n (%) | $\chi^2$ (p-value) | Good perception (n = 1,276) | | |
| --- | --- | --- | --- | --- | --- | --- |
|  |  |  |  | p-value | OR (95% C.I) | aOR (95% C.I) |
| **1.** | **Age (in years)** |  |  |  |  |  |
| | 31 – 40/ $\leq 30$ | 548/ 678 | 6.331 (**$4.2 \times 10^{-2}$**) | $6.7 \times 10^{-1}$ | 1.0 (0.8 – 1.2) | 1.0 (0.8 – 1.2) |
| | $\geq 41/ \leq 30$ | 50 (3.9) | | $2.1 \times 10^{-1}$ | †1.3 (0.8 – 2.2) | †1.2 (0.8 – 1.7) |
| **2.** | **Religion** |  |  |  |  |  |
|  | Protestant/Catholic | 1,202/ 74 | 21.907 (**$2.9 \times 10^{-6}$**) | **$1.7 \times 10^{-2}$** | †2.4 (1.2 – 4.8) | †2.5 (1.6 – 3.9) |
| **3.** | **Marital status** |  |  |  |  |  |
| | Single/Married | 222/ 996 | 6.437 ($9.2 \times 10^{-2}$) | $6.2 \times 10^{-1}$ | 1.0 (0.7 – 1.3) | 1.0 (0.8 – 1.1) |
| | Divorced/Married | 44 (3.4) | | $9.3 \times 10^{-1}$ | 0.6 (0.3 – 1.3) | †1.1 (0.7 – 1.5) |
|  | Widowed/Married | 14 (1.1) |  | **$2.9 \times 10^{-3}$** | 0.7 (0.3 – 1.6) | 0.6 (0.3 – 0.9) |
| **4.** | **Education** |  |  |  |  |  |
| | Secondary/Tertiary | 644/ 71 | 231.220 (**$< 0.001$**) | $6.9 \times 10^{-1}$ | †1.3 (0.6 – 3.0) | 0.9 (0.5 – 1.5) |
|  | Primary/Tertiary | 494 (38.7) |  | **$6.3 \times 10^{-3}$** | †3.1 (1.3 – 6.9) | †2.1 (1.2 – 3.6) |
|  | NFE/Tertiary | 67 (5.3) |  | **$1.0 \times 10^{-4}$** | †5.7 (2.3 – 13.7) | †5.1 (2.8 – 8.9) |
| **5.** | **# of children [2 - 4/ 1]** | 50/ 1,226 | 0.252 ($6.2 \times 10^{-1}$) | $4.2 \times 10^{-1}$ | 0.9 (0.5 – 1.7) | 0.8 (0.5 – 1.3) |
| **6.** | **Occupation** |  |  |  |  |  |
|  | Artisan/Civil Servant | 477/ 79 | 486.781 (**$< 0.001$**) | **$1.0 \times 10^{-4}$** | 0.8 (0.5 – 1.6) | 0.5 (0.3 – 0.7) |
|  | Casual labour/Civil Servant | 172 (13.5) |  | **$< 0.001$** | 0.2 (0.1 – 0.5) | 0.2 (0.1 – 0.3) |
|  | Unemployed/Civil Servant | 548 (42.9) |  | **$< 0.001$** | 0.06 (0.03 – 0.1) | 0.03 (0.02 – 0.04) |
| **7.** | **AMI (in \$)** | | | | | |
| | 101 – 200/ $< 100$ | 138/ 878 | 81.611 (**$< 0.001$**) | **$2.5 \times 10^{-2}$** | 0.6 (0.4 – 0.9) | 0.4 (0.3 – 0.5) |
| | 201 – 300/ $< 100$ | 135 (10.6) | | **$< 0.001$** | 0.3 (0.2 – 0.5) | 0.1 (0.08 – 0.2) |
| | $> 400/ < 100$ | 125 (9.8) | | **$< 0.001$** | 0.3 (0.2 – 0.5) | 0.05 (0.03 – 0.09) |
|  | CONSTANT |  |  | **$6.5 \times 10^{-2}$** |  |  |

**Legend.** #; number, %; proportion of respondents, †; Most likely category, 95% C.I; 95% Confidence interval, AMI; Average Monthly Income, aOR; Adjusted odds ratio, **Boldface** numbers indicate significant p-values, n; frequency/count, NFE; No Formal Education, OR; Odds ratio, Reference Category; Good perception, $\chi^2$ ; Pearson Chi-Square.

### Vaccine misinformation

Generally, 379 (18.8%, 95% C.I; 17.2 – 20.6) of the mothers respectively admitted that messages from anti-vaccine groups affect their confidence in vaccinating their children.

Logistic regression analysis revealed that age, marital status, and monthly income of the mothers were significantly associated with, misinformation concerning vaccines. Age, marital status, and monthly income were also found to be significantly associated with trust in misinformation of vaccines. From multinomial regression analysis, the odds for being misinformed about vaccines was higher amongst protestants (*p* = 1.0×10^−5^, aOR; 1.7, 95%C.I; 1.4 – 2.3), secondary school leavers (*p* = 2.6×10^−1^, aOR; 1.2, 95%C.I; 0.8 – 1.8), primary school leavers (*p* = 8.2×10^−1^, aOR; 1.0, 95%C.I; 0.7 – 1.8), and mothers with NFE (*p* = 8.5×10^−1^, aOR; 1.0, 95%C.I; 0.6 – 1.6), when compared with their other counterparts. The odds for trusting vaccine misinformation were higher amongst protestants (*p* = 7.1×10^−1^, aOR; 1.2, 95%C.I; 0.5 – 2.5) when compared with the catholic category (Table 5).

**Table 5:** Multivariable logistic regression of demographic predictors of anti-vaccine misinformation vs trust of misinformation.

| S/N | Characteristic | Anti-vaccine misinformation (n = 379) |  |  |  | Trust of misinformation (n = 1,968) |  |  |  |
| --- | --- | --- | --- | --- | --- | --- | --- | --- | --- |
|  |  | n (%) | p-value | OR (95% C.I) | aOR (95% C.I) | n (%) | p-value | OR (95% C.I) | aOR (95% C.I) |
| **1.** | **Age (in years)** |  |  |  |  |  |  |  |  |
|  | 31 – 40/ ≤ 30 | 173/ 181 | **2.6x10<sup>-3</sup>** | 0.8 (0.6 – 1.0) | 0.8 (0.7 – 0.9) | 845/ 1,027 | **3.7x10<sup>-3</sup>** | 0.7 (0.4 – 1.1) | 0.6 (0.5 – 0.8) |
|  | ≥ 41/ ≤ 30 | 25 (6.6) | **1.0x10<sup>-5</sup>** | 0.5 (0.3 – 0.9) | 0.4 (0.3 – 0.5) | 96 (4.9) | 1.6x10<sup>-1</sup> | 0.5 (0.2 – 1.9) | 0.5 (0.3 – 1.2) |
| **2.** | **Religion** |  |  |  |  |  |  |  |  |
|  | Protestant/Catholic | 353/ 26 | **1.0x10<sup>-5</sup>** | †1.9 (1.2 – 3.1) | †1.7 (1.4 – 2.3) | 1,885/ 83 | 7.1x10<sup>-1</sup> | †1.1 (0.3 – 5.0) | †1.2 (0.5 – 2.5) |
| **3.** | **Marital status** |  |  |  |  |  |  |  |  |
|  | Single/Married | 63/ 291 | 2.7x10<sup>-1</sup> | 0.8 (0.6 – 1.2) | 0.9 (0.7 – 1.1) | 361/ 1,522 | **2.6x10<sup>-2</sup>** | 0.6 (0.3 – 1.2) | 0.6 (0.4 – 0.9) |
|  | Divorced/Married | 18 (4.7) | **8.6x10<sup>-3</sup>** | 0.5 (0.3 – 1.0) | 0.6 (0.5 – 0.9) | 59 (3.0) | 9.5x10<sup>-1</sup> | 0.03 (0.01 – 0.05) | 0.03 (0.02 – 0.05) |
|  | Widowed/Married | 7 (1.8) | 1.1x10<sup>-1</sup> | 0.6 (0.2 – 1.5) | 0.6 (0.4 – 1.1) | 26 (1.3) | 1.8x10<sup>-1</sup> | 0.6 (0.07 – 4.7) | 0.3 (0.09 – 1.6) |
| **4.** | **Education** |  |  |  |  |  |  |  |  |
|  | Secondary/ Tertiary | 158/ 17 | 2.6x10<sup>-1</sup> | †1.3 (0.6 – 2.6) | †1.2 (0.8 – 1.8) | 785/ 82 | 3.2x10<sup>-1</sup> | †1.2 (0.1 – 12.8) | 0.5 (0.1 – 2.0) |
|  | Primary/ Tertiary | 173 (45.6) | 8.2x10<sup>-1</sup> | †1.5 (0.7 – 3.1) | †1.0 (0.7 – 1.8) | 923 (46.9) | 4.2x10<sup>-1</sup> | †1.2 (0.1 – 13.3) | 0.6 (0.1 – 2.3) |
|  | NFE/ Tertiary | 31 (8.2) | 8.5x10<sup>-1</sup> | †1.7 (0.7 – 3.9) | †1.0 (0.6 – 1.6) | 178 (9.0) | 9.5x10<sup>-1</sup> | †1.8 (0.2 – 21.5) | 0.9 (0.2 – 4.1) |
| **5.** | **# of children [2 - 4/ 1]** | 17/ 362 | 1.3x10<sup>-1</sup> | 0.8 (0.4 – 1.4) | 0.8 (0.6 – 1.1) | 76/ 1,892 | 9.6x10<sup>-1</sup> | 0.05 (0.03 – 2.1) | 0.05 (0.03 – 2.0) |
| **6.** | **Occupation** |  |  |  |  |  |  |  |  |
|  | Artisan/ Civil Servant | 228/ 11 | **1.0x10<sup>-4</sup>** | 0.2 (0.08 – 0.4) | 0.1 (0.07 – 0.2) | 1,082/ 106 | 7.9x10<sup>-2</sup> | 0.4 (0.1 – 1.5) | 0.5 (0.2 – 1.0) |
|  | Casual labour/ Civil Servant | 59 (15.6) | **1.0x10<sup>-5</sup>** | 0.2 (0.07 – 0.3) | 0.2 (0.1 – 0.3) | 190 (9.6) | 9.5x10<sup>-2</sup> | 0.3 (0.05 – 1.9) | 0.4 (0.1 – 1.2) |
|  | Unemployed/ Civil Servant | 81 (21.4) | **1.0x10<sup>-5</sup>** | 0.3 (0.1 – 0.7) | 0.2 (0.1 – 0.4) | 590 (29.9) | **2.2x10<sup>-5</sup>** | 0.1 (0.04 – 0.4) | 0.09 (0.04 – 0.2) |
| **7.** | **AMI (in \$)** | | | | | | | | |
|  | 101 – 200/ < 100 | 37/ 254 | **1.0x10<sup>-5</sup>** | 0.9 (0.6 – 1.5) | 0.6 (0.4 – 0.7) | 191/ 1,465 | **2.0x10<sup>-4</sup>** | 0.2 (0.04 – 0.9) | 0.2 (0.06 – 0.4) |
|  | 201 – 300/ < 100 | 43 (11.4) | **1.0x10<sup>-5</sup>** | 0.6 (0.3 – 1.0) | 0.4 (0.3 – 0.5) | 166 (8.4) | **3.0x10<sup>-4</sup>** | 0.1 (0.01 – 0.9) | 0.02 (0.01 – 0.1) |
|  | > 400/ < 100 | 45 (11.9) | **3.0x10<sup>-4</sup>** | 0.4 (0.3 – 0.7) | 0.2 (0.1 – 0.3) | 146 (7.2) | **2.0x10<sup>-4</sup>** | 0.1 (0.01 – 1.1) | 0.01 (0.001 – 0.1) |
|  | **CONSTANT** |  | **1.0x10<sup>-5</sup>** |  |  |  | 1.5x10<sup>-1</sup> |  |  |

**Legend.** #, number, %; proportion of respondents, †; Most likelihood category, 95% C.I; 95% Confidence interval, AMI; Average Monthly Income, aOR; Adjusted odds ratio, **Boldface** numbers indicate significant *p*-values, *n*; frequency/count, NFE; No Formal Education, OR; Odds ratio, Reference Category; No.

### Effect of misinformation on the continuity of routine immunisation

On misinformation about COVID-19 vaccines, 1,563 (76.4%, 95% C.I; 74.5 – 78.2) admitted to having received or heard negative information about the vaccines.

Logistic regression analysis revealed that except for age, all the demographic characteristics of respondents were significantly associated with, COVID-19 misinformation. Age, marital status, and monthly income were also found to be significantly associated with the trust of COVID-19 misinformation and the effect of COVID-19 on RI (Table 6).

**Table 6:**
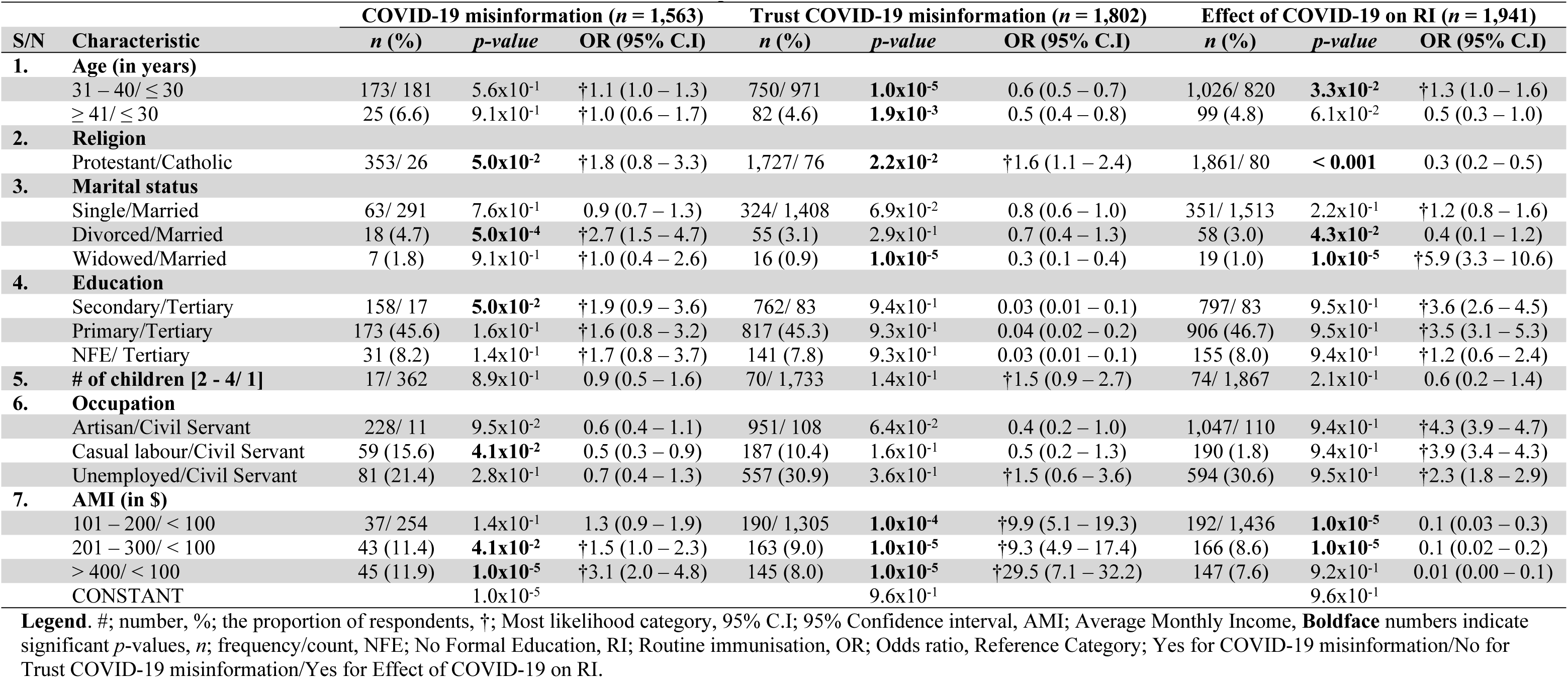
Misinformation, trust in misinformation and effect of the COVID-19 pandemic on RI.

| S/N | Characteristic | COVID-19 misinformation (*n* = 1,563) |  |  | Trust COVID-19 misinformation (*n* = 1,802) |  |  | Effect of COVID-19 on RI (*n* = 1,941) |  |  |
| --- | --- | --- | --- | --- | --- | --- | --- | --- | --- | --- |
|  |  | *n* (%) | *p*-value | OR (95% C.I) | *n* (%) | *p*-value | OR (95% C.I) | *n* (%) | *p*-value | OR (95% C.I) |
| **1.** | **Age (in years)** |  |  |  |  |  |  |  |  |  |
|  | 31 – 40/ ≤ 30 | 173/ 181 | 5.6x10<sup>-1</sup> | †1.1 (1.0 – 1.3) | 750/ 971 | **1.0x10<sup>-5</sup>** | 0.6 (0.5 – 0.7) | 1,026/ 820 | **3.3x10<sup>-2</sup>** | †1.3 (1.0 – 1.6) |
|  | ≥ 41/ ≤ 30 | 25 (6.6) | 9.1x10<sup>-1</sup> | †1.0 (0.6 – 1.7) | 82 (4.6) | **1.9x10<sup>-3</sup>** | 0.5 (0.4 – 0.8) | 99 (4.8) | 6.1x10<sup>-2</sup> | 0.5 (0.3 – 1.0) |
| **2.** | **Religion** |  |  |  |  |  |  |  |  |  |
|  | Protestant/Catholic | 353/ 26 | **5.0x10<sup>-2</sup>** | †1.8 (0.8 – 3.3) | 1,727/ 76 | **2.2x10<sup>-2</sup>** | †1.6 (1.1 – 2.4) | 1,861/ 80 | **< 0.001** | 0.3 (0.2 – 0.5) |
| **3.** | **Marital status** |  |  |  |  |  |  |  |  |  |
|  | Single/Married | 63/ 291 | 7.6x10<sup>-1</sup> | 0.9 (0.7 – 1.3) | 324/ 1,408 | 6.9x10<sup>-2</sup> | 0.8 (0.6 – 1.0) | 351/ 1,513 | 2.2x10<sup>-1</sup> | †1.2 (0.8 – 1.6) |
|  | Divorced/Married | 18 (4.7) | **5.0x10<sup>-4</sup>** | †2.7 (1.5 – 4.7) | 55 (3.1) | 2.9x10<sup>-1</sup> | 0.7 (0.4 – 1.3) | 58 (3.0) | **4.3x10<sup>-2</sup>** | 0.4 (0.1 – 1.2) |
|  | Widowed/Married | 7 (1.8) | 9.1x10<sup>-1</sup> | †1.0 (0.4 – 2.6) | 16 (0.9) | **1.0x10<sup>-5</sup>** | 0.3 (0.1 – 0.4) | 19 (1.0) | **1.0x10<sup>-5</sup>** | †5.9 (3.3 – 10.6) |
| **4.** | **Education** |  |  |  |  |  |  |  |  |  |
|  | Secondary/Tertiary | 158/ 17 | **5.0x10<sup>-2</sup>** | †1.9 (0.9 – 3.6) | 762/ 83 | 9.4x10<sup>-1</sup> | 0.03 (0.01 – 0.1) | 797/ 83 | 9.5x10<sup>-1</sup> | †3.6 (2.6 – 4.5) |
|  | Primary/Tertiary | 173 (45.6) | 1.6x10<sup>-1</sup> | †1.6 (0.8 – 3.2) | 817 (45.3) | 9.3x10<sup>-1</sup> | 0.04 (0.02 – 0.2) | 906 (46.7) | 9.5x10<sup>-1</sup> | †3.5 (3.1 – 5.3) |
|  | NFE/ Tertiary | 31 (8.2) | 1.4x10<sup>-1</sup> | †1.7 (0.8 – 3.7) | 141 (7.8) | 9.3x10<sup>-1</sup> | 0.03 (0.01 – 0.1) | 155 (8.0) | 9.4x10<sup>-1</sup> | †1.2 (0.6 – 2.4) |
| **5.** | **# of children [2 - 4/ 1]** | 17/ 362 | 8.9x10<sup>-1</sup> | 0.9 (0.5 – 1.6) | 70/ 1,733 | 1.4x10<sup>-1</sup> | †1.5 (0.9 – 2.7) | 74/ 1,867 | 2.1x10<sup>-1</sup> | 0.6 (0.2 – 1.4) |
| **6.** | **Occupation** |  |  |  |  |  |  |  |  |  |
|  | Artisan/Civil Servant | 228/ 11 | 9.5x10<sup>-2</sup> | 0.6 (0.4 – 1.1) | 951/ 108 | 6.4x10<sup>-2</sup> | 0.4 (0.2 – 1.0) | 1,047/ 110 | 9.4x10<sup>-1</sup> | †4.3 (3.9 – 4.7) |
|  | Casual labour/Civil Servant | 59 (15.6) | **4.1x10<sup>-2</sup>** | 0.5 (0.3 – 0.9) | 187 (10.4) | 1.6x10<sup>-1</sup> | 0.5 (0.2 – 1.3) | 190 (1.8) | 9.4x10<sup>-1</sup> | †3.9 (3.4 – 4.3) |
|  | Unemployed/Civil Servant | 81 (21.4) | 2.8x10<sup>-1</sup> | 0.7 (0.4 – 1.3) | 557 (30.9) | 3.6x10<sup>-1</sup> | †1.5 (0.6 – 3.6) | 594 (30.6) | 9.5x10<sup>-1</sup> | †2.3 (1.8 – 2.9) |
| **7.** | **AMI (in \$)** | | | | | | | | | |
|  | 101 – 200/ < 100 | 37/ 254 | 1.4x10<sup>-1</sup> | 1.3 (0.9 – 1.9) | 190/ 1,305 | **1.0x10<sup>-4</sup>** | †9.9 (5.1 – 19.3) | 192/ 1,436 | **1.0x10<sup>-5</sup>** | 0.1 (0.03 – 0.3) |
|  | 201 – 300/ < 100 | 43 (11.4) | **4.1x10<sup>-2</sup>** | †1.5 (1.0 – 2.3) | 163 (9.0) | **1.0x10<sup>-5</sup>** | †9.3 (4.9 – 17.4) | 166 (8.6) | **1.0x10<sup>-5</sup>** | 0.1 (0.02 – 0.2) |
|  | > 400/ < 100 | 45 (11.9) | **1.0x10<sup>-5</sup>** | †3.1 (2.0 – 4.8) | 145 (8.0) | **1.0x10<sup>-5</sup>** | †29.5 (7.1 – 32.2) | 147 (7.6) | 9.2x10<sup>-1</sup> | 0.01 (0.00 – 0.1) |
|  | CONSTANT |  | 1.0x10<sup>-5</sup> |  |  | 9.6x10<sup>-1</sup> |  |  | 9.6x10<sup>-1</sup> |  |

**Legend.** #; number, %; the proportion of respondents, †; Most likelihood category, 95% C.I; 95% Confidence interval, AMI; Average Monthly Income, **Boldface** numbers indicate significant *p*-values, *n*; frequency/count, NFE; No Formal Education, RI; Routine immunisation, OR; Odds ratio, Reference Category; Yes for COVID-19 misinformation/No for Trust COVID-19 misinformation/Yes for Effect of COVID-19 on RI.

The factors affecting routine immunisation are presented in Table 7. Logistic regression analysis revealed that all the factors explored were significantly associated with RI. Culture, lack of funds hindering immunisation, trusting in misinformation, disagreeing on serious adverse effects of vaccines as well as disagreeing on serious adverse effects of COVID-19 vaccines were respectively; about seven times (6.8, 95% C.I:4.6 – 10.2), one and a half times (1.5, 95% C.I: 1.1 – 2.1), two and a half times (2.5, 95% C.I: 1.6 – 3.9), about four times (3.8, 9.5% C.I: 2.8 – 5.1) and about one and a third times (1.6, 95% C.I; 1.2 – 2.2) more likely to affect RI when compared with their various counterparts.

**Table 7:** Factors affecting RI.

| S/N | Characteristic | Subclass | Effect of COVID-19 on RI (n = 1,941) |  |  |  |
| --- | --- | --- | --- | --- | --- | --- |
|  |  |  | Yes (%) | No (%) | p-value | OR (95% C.I) |
| 1. | Culture supports immunisation | Yes | 87 (83.6) | 1,786 (92.0) | < **0.001** | †6.8 (4.6 – 10.2) |
|  |  | No | 17 (16.4) | 155 (8.0) |  |  |
| 2. | Lack of funds hinders immunisation | Yes | 69 (66.4) | 1,778 (91.6) | **1.1x10<sup>-2</sup>** | †1.5 (1.1 – 2.1) |
|  |  | No | 35 (33.6) | 163 (8.4) |  |  |
| 3. | Does vaccine misinformation affect your vaccine confidence?* | Yes | 62 (62.0) | 317 (16.6) | < **0.001** | 0.1 (0.1 – 0.2) |
|  |  | No | 38 (38.0) | 1,597 (83.4) |  |  |
| 4. | Do you trust vaccine misinformation? | Yes | 93 (89.4) | 1,875 (96.6) | < **0.001** | †2.5 (1.6 – 3.9) |
|  |  | No | 11 (10.6) | 66 (3.4) |  |  |
| 5. | Concerned about adverse effects of vaccines | Disagree | 43 (41.4) | 422 (21.7) | < **0.001** | †3.8 (2.8 – 5.1) |
|  |  | Agree | 61 (58.6) | 1,519 (78.3) |  |  |
| 6. | Concerned about adverse effects of COVID-19 vaccines | Disagree | 42 (40.4) | 802 (41.3) | **1.2x10<sup>-3</sup>** | †1.6 (1.2 – 2.2) |
|  |  | Agree | 62 (59.6) | 1,139 (58.7) |  |  |
| 7. | Concerned that COVID-19 vaccines might not be safe | Disagree | 60 (57.7) | 1,819 (93.7) | < **0.001** | 0.09 (0.07 – 0.1) |
|  |  | Agree | 44 (43.3) | 122 (6.3) |  |  |
|  |  | Total | 104 | 1,941 |  |  |

**Legend.** %; proportion of respondents, †; Most likely category, 95% C.I; 95% Confidence interval, RI; Routine immunisation

Of the 1,941 mothers who said the COVID-19 pandemic affected RI, 1,786 (92.0%), 1,778 (91.6%), 1,875 (96.6%), and 1,519 (78.3%) admitted that culture supports immunisation, the lack of funds to visit the health facilities hinders immunisation, trusted vaccine misinformation, and had concerns about the adverse effects of vaccines respectively. Regarding the adverse effects of vaccines and COVID-19 vaccines, the mothers were about four times (OR; 3.8, 95% CI: 2.8 – 5.1), and one and a half times (OR; 1.6, 95% CI: 1.2 – 2.2) more likely to disagree that they are concerned with the serious adverse effects of vaccines in general and COVID-19 vaccines in particular when compared with having to agree on these adverse effects (Table 7).

## Discussion

Our study adds to the description of vaccine continuation and completion in Rwanda. This study revealed that; 94.1% of mothers obtained information about vaccines from health care workers, 5.5% had poor access to vaccines, 9.7% complained of lack of transport fare to the vaccination centre, 62.4% had a good perception of vaccine risks, 76.4% had been misinformed on COVID-19 vaccines, 96.2% trust vaccine misinformation, 88.1% trust COVID-19 vaccine misinformation, and 94.9% admitted that RI of their children was affected by the COVID-19 pandemic.

### Sources of information

We observed that the major source of information on vaccines and vaccination was provided by healthcare providers (94.1%), followed by the mass media and social media (28.8%) and relationships (7.1%). This was in line with previous studies in Nigeria, Cyprus and Switzerland, where parents rely on paediatricians and nurses for information concerning childhood vaccination[50–52], but differed from studies in the Netherlands, Philippines, and Guinea where parents mostly explored the internet, as well as relied on traditional authorities for information about childhood vaccinations[53–55]. In other studies, parents considered factors like access to information, interpersonal communication, misinformation, and community norms for childhood vaccination[56–59]. The differences in the rates and variety of sources of vaccine information could be explained by the fact that the present study was a nationwide-based study in which many sources of information on vaccines were explored amongst mothers.

### Reasons for vaccination/immunisation discontinuation

The 5.5% proportion of mothers who complained of inaccessibility to healthcare facilities in this study was low compared with a study on the concerns about COVID-19 accessibility, affordability, and acceptability[60]. Our finding of inaccessibility to healthcare facilities was different from; perceived risk of disease, vaccine efficacy, prior negative experience, side effects and the social environment as reported in focus group studies in Kenya and the Netherlands[61,62].

### Religious and traditional tendencies towards the immunisation of children

In our study, 92.2% and 91.6% of the mothers indicated that their religion and culture were in support of childhood vaccination and immunisation. In our study, religion was associated with age, marital status, education and average monthly income; which was synonymous with maternal education being associated with vaccination coverage in 127 countries worldwide[63]. The support of childhood immunisation by culture is in line with ethnic backgrounds as reported amongst Black and Asian minority ethnic groups in the United Kingdom[64].

### Mothers’ perception of risks

In our study, 77.3%, 58.7%, and 8.1% of the mothers respectively agreed to the fact that; there were serious AEs of vaccines, serious AEs of COVID-19 vaccines, and that the COVID-19 vaccines may not be safe. This was similar to the fear of vaccines, expressed by mothers in a study reported in Australia[65], the fear of COVID-19 vaccines due to history of COVID-19 infection in Nepal[66], and contrary to the perception of 91.7% of Greek mothers who believed that COVID-19 vaccines would protect their children[67]. The 91.9% rate of respondents in our study who agree that COVID-19 vaccines may be safe was less than the 45.3% reported among parents in Egypt[68]. The differences in the perception of either vaccine or COVID-19 vaccine risks may be due to the differences in the study populations as well as study designs.

### Vaccine misinformation

In this study, 18.5% of the mothers had been misinformed about vaccines in general, and 76.4% had been misinformed about COVID-19 vaccines. The 76.4% misinformation on COVID-19 vaccine in this study is more than the 57.6% in the USA[69]. Our study revealed that age, religion, marital status, occupation and monthly income of the mothers were significantly associated with, misinformation concerning vaccines.

### Factors Affecting Routine Immunisation

The 94.9% who admitted that RI of their children, were synonymous with rates reported worldwide[70] as well as the 96.3% of measles immunisation post-coverage reported in Nigeria[71]. This was however higher when compared with the 72% rate for DPT3 reported in Laos[72]; the 71 – 72% Measles-Rubella and yellow fever virus, 71% OPV3, 82.5% Penta 3, and 83.5% BCG reported in the rural settings of the Gambia and India[73,74], as well as the 18.8 – 69.1% reported among Pakistani children[75]. The differences may be due to differences in study designs.

In our study, the following factors affected RI the lack of funds, COVID-19 vaccine misinformation, concern about the adverse effects of vaccines and COVID-19 vaccines, as well as the safety of COVID-19 vaccines. Our findings were different from; the willingness of mothers to pay for immunisation, distance to the immunisation, illiteracy, occupation, family size, home delivery and health education, as reported in Laos as well as the Gambia[72,73]. Our findings were however similar to the educational level of parents, occupation and source of healthcare information among Pakistani parents[75].

### Strengths and limitations

#### Strengths

The data collection was carried out by field staff who were familia with the local terrain, ensuring a thorough understanding of the study areas. The quality of the data collected was assured through pretesting of questionnaires in a pilot study. The pilot study aimed to minimise bias and errors, improving the accuracy and reliability of the data. The minimisation of bias was done by randomisation in the selection of Provinces, Districts, Sectors, Cells, and Villages, screening and excluding other caregivers who were not mothers, as well as sorting out and eliminating outliers.

#### Limitations

This was a cross-sectional study, representing the snapshot of the population within the study period. Thus, we cannot infer causal relationships between mothers’ religion/culture, perception of risks, and continuity of childhood vaccination with their demographic characteristics. Data were collected through anonymous self-reporting via door-to-door, and thus there is a possibility of response bias and recall bias. Such biases can also affect some of the responses as well as the results of the study. Another significant limitation was representativeness as a higher proportion of the sampled respondents were undereducated, having acquired less than the secondary level of education.

## Conclusion

During the COVID-19 pandemic, there was an interruption of routine immunisation worldwide. In the case of Rwanda, the impact of COVID-19 on routine immunisation was not significantly felt due to Rwanda’s initial preparedness to safeguard against infectious diseases such as Ebola. This study indicates that there was an association between RI with culture, lack of funds, trust in vaccine misinformation, concern about adverse effects of vaccines and COVID-19 vaccines, as well as the safety of the COVID-19 vaccines.

## Abbreviations

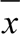: Mean
95%C.I: 95%Confidence Interval
AEs: Adverse effects
aOR: Adjusted Odds Ratio
BCG: Bacille Calmette-Guérin
COVID-19: Coronavirus Disease 2019
DTP3: 3^rd^ dose Diphtheria Tetanus Toxoid and Pertussis
MCV1: 1^st^ dose of Measles Containing Vaccine
OPV3: 3^rd^ dose of Oral Poliovirus Vaccine
OR: Odds Ratio
RI: Routine Immunisation
VH: Vaccine hesitancy
χ^2^: Pearson Chi square.

## Supplementary Information

**S1 Appendix**. Survey Questionnaire (DOC)

**S2 Appendix**. Supplementary Tables (DOC)

## Declarations

### Ethics approval to participate

This study was conducted in accordance with the Declaration of Helsinki and approved by the Institutional Review Board (IRB) of the University of Rwanda, College of Medicine and Health Science (No. 402/CMHS IRB/2020). All participants signed the informed consent prior to being interviewed.

### Consent for publication

Not applicable

### Data availability statement

All relevant data that support the conclusion of this study are included in the article.

### Competing interests

The authors disclose no conflicts of interest.

### Funding

This study was funded by the Government of Rwanda and the German Academic Exchange Service (DAAD) with Funding ID 57398664.

### Author Contributions

**Conceptualisation**: Edward Mbonigaba, Claudia M Denkinger, Shannon A McMahon, and Simiao Chen.

**Supervision**: Shannon A McMahon, Claudia M Denkinger, Simiao Chen.

**Data curation & Formal analysis**: Edward Mbonigaba, Mark Donald C Reñosa, Frederick Nchang Cho.

**Investigation**: Edward Mbonigaba.

**Methodology**: Edward Mbonigaba, Fengyun Yu, Frederick Nchang Cho, Wenjin Chen and Simiao Chen.

**Project administration**: Edward Mbonigaba, Mark Donald C. Reñosa, Claudia M. Denkinger, and Simiao Chen.

**Resources**: Edward Mbonigaba, Claudia M. Denkinger, and Simiao Chen.

**Validation**: Edward Mbonigaba, Fengyun Yu, Mark Donald C. Reñosa, Frederick Nchang Cho, Qiushi Chen, Wenjin Chen,Shannon A McMahon, Claudia M. Denkinger, and Simiao Chen.

**Writing – Original draft**: Edward Mbonigaba, Frederick Nchang Cho, Shannon A McMahon, Claudia M. Denkinger, and Simiao Chen.

**Writing – review and editing**: Edward Mbonigaba, Fengyun Yu, Mark Donald C. Reñosa, Frederick Nchang Cho, Shannon A McMahon, Claudia M. Denkinger, and Simiao Chen.

## Acknowledgements

The authors gratefully acknowledge the Government of Rwanda and the German Academic Exchange Service (DAAD) for providing grant funding (Funding -ID 57398664) for this study.

The views expressed in this research article are those of the authors and may not necessarily reflect those of their affiliated institutions.

## Notes

### Competing Interest Statement

The authors have declared no competing interest.

### Funding Statement

This study was funded by the Government of Rwanda and the German Government through Germany Academic Exchange Service

### Author Declarations

Institutional Review Board (IRB) of the University of Rwanda, College of Medicine and Health Science (No. 402/CMHS IRB/2020)

